# Potential biases arising from epidemic dynamics in observational seroprotection studies

**DOI:** 10.1101/2020.05.02.20088765

**Authors:** Rebecca Kahn, Lee Kennedy-Shaffer, Yonatan H. Grad, James M. Robins, Marc Lipsitch

## Abstract

The extent and duration of immunity following SARS-CoV-2 infection are critical outstanding questions about the epidemiology of this novel virus, and studies are needed to evaluate the effects of serostatus on reinfection. Understanding the potential sources of bias and methods to alleviate biases in these studies is important for informing their design and analysis. Confounding by individual-level risk factors in observational studies like these is relatively well appreciated. Here, we show how geographic structure and the underlying, natural dynamics of epidemics can also induce noncausal associations. We take the approach of simulating serologic studies in the context of an uncontrolled or a controlled epidemic, under different assumptions about whether prior infection does or does not protect an individual against subsequent infection, and using various designs and analytic approaches to analyze the simulated data. We find that in studies assessing the efficacy of serostatus on future infection, comparing seropositive individuals to seronegative individuals with similar time-dependent patterns of exposure to infection, by stratifying or matching on geographic location and time of enrollment, is essential to prevent bias.

---

The extent and duration of immunity following SARS-CoV-2 infection are critical outstanding questions about the epidemiology of this novel virus (1). Serologic tests, which detect the presence of antibodies, are becoming more widely available (2). However, the presence of antibodies, or seroconversion, does not guarantee immunity to reinfection, and experimental data with other coronaviruses raise concerns that antibodies could under some circumstances enhance future infections (3). Studies are needed to evaluate the short and long term effects of seropositivity. Understanding the potential sources of bias and methods to alleviate biases in these studies is important for informing their design and analysis.

We consider observational studies, in which the exposure (prior infection or seropositivity) is not assigned at random. Such studies always face the risk of confounding: factors that directly influence both the exposure and (separately) the outcome. Studies of seropositivity and its effect on future infection are particularly prone to confounding because the exposure (a marker of previous infection) and the outcome (future infection) are almost the same event in the same person, separated in time. Thus factors that influence the risk of infection are nearly always potential confounders. For example, individuals in high-risk occupations (e.g., health care workers) are more likely to become seropositive and are more likely to be exposed again once they are seropositive. Confounding by individual-level risk factors is relatively well appreciated. Less obvious perhaps is that geographic structure (4) or the underlying, natural dynamics of epidemics (5,6) can induce noncausal associations between an exposure and an outcome. For example, if the overall size of an epidemic is very different in different communities, individuals in communities with small epidemics will have low prevalence of the exposure (seropositivity) and low incidence of the outcome (infection after enrollment). If individuals are enrolled at different times during an epidemic with an upward trajectory (such as the early exponential phase of an epidemic), individuals enrolled early in the epidemic will be less likely to be seropositive (exposure) and less likely to become infected at a given point in time after enrollment (outcome) than those with a later date of enrollment. In an epidemic that is controlled (thus with an up-then-down trajectory of incidence) the representation of seropositive individuals will increase with time, but the rate at which these individuals experience the outcome will increase then decrease, creating potential for confounding in either direction.

In this study we take the approach of simulating such studies in the context of an uncontrolled or a controlled epidemic, under different assumptions about whether prior infection does or does not protect an individual against subsequent infection, and using various designs and analytic approaches to analyze the simulated data. By identifying the direction and comparative magnitude of bias of the estimated degree of protection relative to a known true effect of prior infection (known because we have built it into the simulations), we identify means of designing and analyzing such studies that can render them less likely to show bias due to these confounding factors. This framework of simulating trials in the context of an epidemic has been widely used to understand experimental (7) and observational (5,8) studies of risk factors and prevention interventions for infectious disease.

## METHODS

We simulate a stochastic outbreak of a disease in a network of people grouped into communities, with each community’s outbreak seeded by introductions over time (4,9). For each simulation, we generate a network graph, where individuals are grouped into either one community of 10,000 people or 10 communities of 1,000 people each. People are only connected to individuals in their own community, with the probability of such a connection based on an input parameter in the simulation. For “well mixed” communities, every individual is connected to every other within their community, while for simulations with “clustered” communities, individuals have a limited number of connections within their community, which creates smaller sub-communities, or “clusters”, by chance. In these latter simulations, individuals may have varying numbers of actual connections but all have the same expected number. The network graph of a “well mixed” community is a complete graph, while that of a “clustered” community is a random graph with uniform edge probability. In simulations with 10 communities, all communities are independent of one another, conditional on the introduction of infection from the outside. At each time step in the model, each susceptible individual has a daily probability of infection from each of their infectious contacts of 1 − *e*^−^*^β^*, where *β* is the force of infection. The outbreak is seeded with stochastic introductions into the communities between days one and fifty based on an external force of infection, which means in simulations with multiple communities, outbreaks may start at different times in each community, and some communities may avoid infection completely.

The disease natural history follows a Susceptible-Exposed-Infectious-Susceptible’ (SEIS’) model, where under the null hypothesis (i.e. no immunity) those in the S and S’ compartments are equally susceptible, while under the alternative hypothesis, those in S’ are less susceptible (in principle, perhaps completely immune, but in keeping with prior evidence about coronaviruses, we assume partially immune) (10,11). In simulations with partial immunity, we make the simplifying assumption that susceptibility is immediately decreased following the infectious period and remains constant over time. Seroconversion is assumed to be detectable seven days after the end of the latent period. We simulate scenarios with no control measures in place and scenarios in which control measures that reduce the force of infection per infected individual (*β*) are implemented at day 90 of the study period. Table 1 shows the specific numbers corresponding to these parameters of the simulations, and the appendix describes the generation of the network and outbreak in more detail.

**Table 1.**
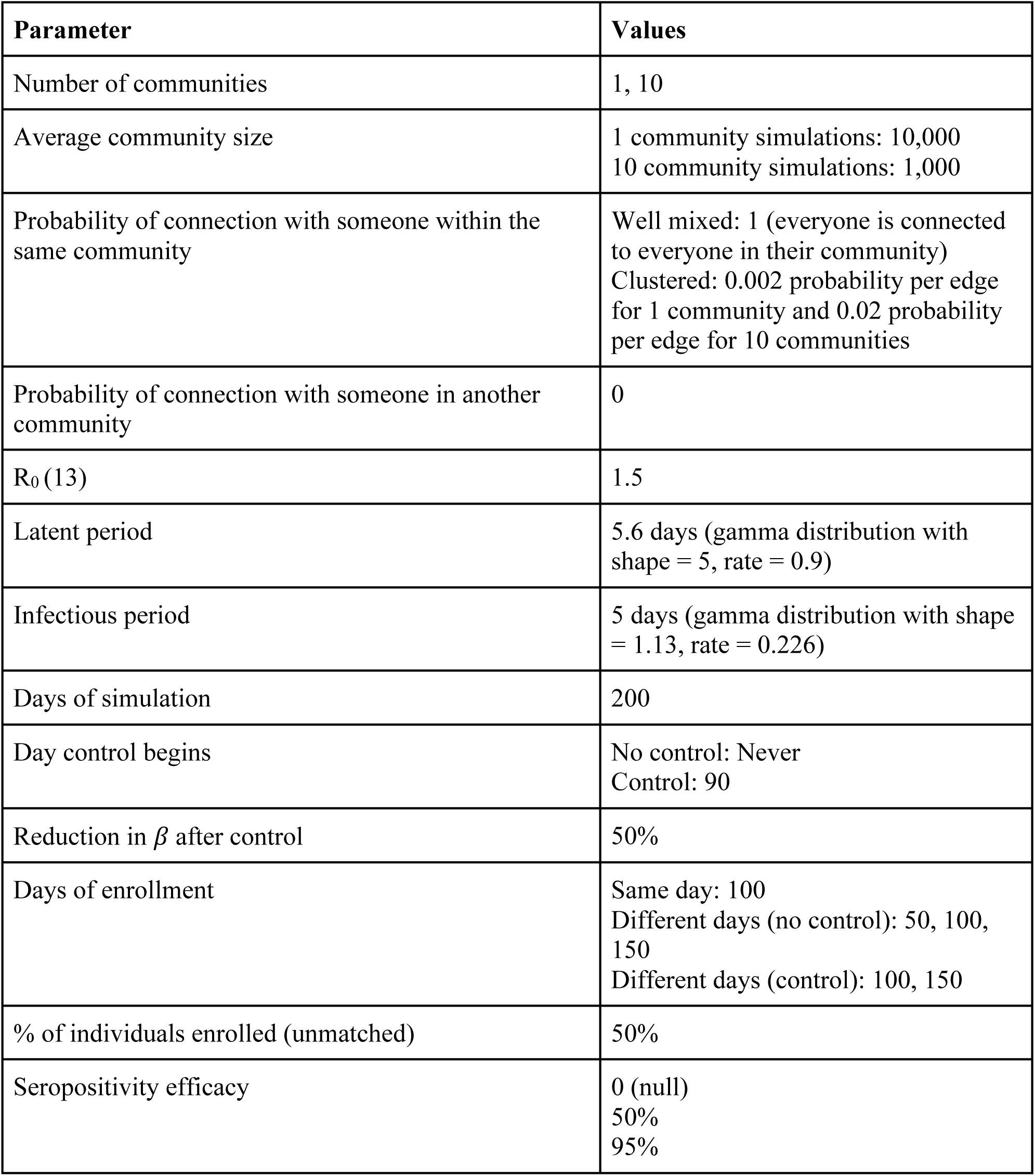

| Parameter | Values |
| --- | --- |
| Number of communities | 1, 10 |
| Average community size | 1 community simulations: 10,000<br>10 community simulations: 1,000 |
| Probability of connection with someone within the same community | Well mixed: 1 (everyone is connected to everyone in their community)<br>Clustered: 0.002 probability per edge for 1 community and 0.02 probability per edge for 10 communities |
| Probability of connection with someone in another community | 0 |
| $R_0$ (13) | 1.5 |
| Latent period | 5.6 days (gamma distribution with shape = 5, rate = 0.9) |
| Infectious period | 5 days (gamma distribution with shape = 1.13, rate = 0.226) |
| Days of simulation | 200 |
| Day control begins | No control: Never<br>Control: 90 |
| Reduction in $\beta$ after control | 50% |
| Days of enrollment | Same day: 100<br>Different days (no control): 50, 100, 150<br>Different days (control): 100, 150 |
| % of individuals enrolled (unmatched) | 50% |
| Seropositivity efficacy | 0 (null)<br>50%<br>95% |

A random sample of individuals are enrolled into the study at specified time points over the course of the outbreak, either all on the same day or at two or three different days throughout the study period. We classify individuals as seropositive or seronegative based on their serostatus on day of enrollment into the study, and then we follow them up until they are infected or until the study period ends at day 200. In the unmatched analyses, we enroll half of the individuals in each community into the study, with an equal number enrolled on each day of enrollment. In the matched analyses, for every seropositive individual enrolled on each day of enrollment, we also enroll one seronegative individual on that day from the same community.

We calculate the hazard ratio of infection comparing seropositive to seronegative individuals, using a Cox proportional hazards model with time starting from enrollment (i.e., possibly not the same calendar time if individuals enroll on different dates). Given the potential for stochasticity to generate heterogeneous outbreaks between communities (4), we compare unstratified analyses to analyses stratified by or matched on community and day of enrollment to prevent confounding by these variables.

## RESULTS

Figure 1 shows the results for 1,000 simulations for each of 36 combinations of parameters. Figures 1A–D summarize results from simulations with no control measures in place, with Figures 1A and 1C under the null, meaning seropositivity has no efficacy against reinfection (*β*^+^ = *β*^−^, where *β*^+^ is the force of infection for contact between an infectious individual and a seropositive individual and *β*^−^ is the force of infection for contact between an infectious individual and a seronegative individual); in Figures B and D, seropositivity reduces susceptibility by 50% *(β^+^* = 0.5\**β^−^*) and 95% *(β^+^* = 0.05\**β^−^*), respectively.

**Figure 1.**
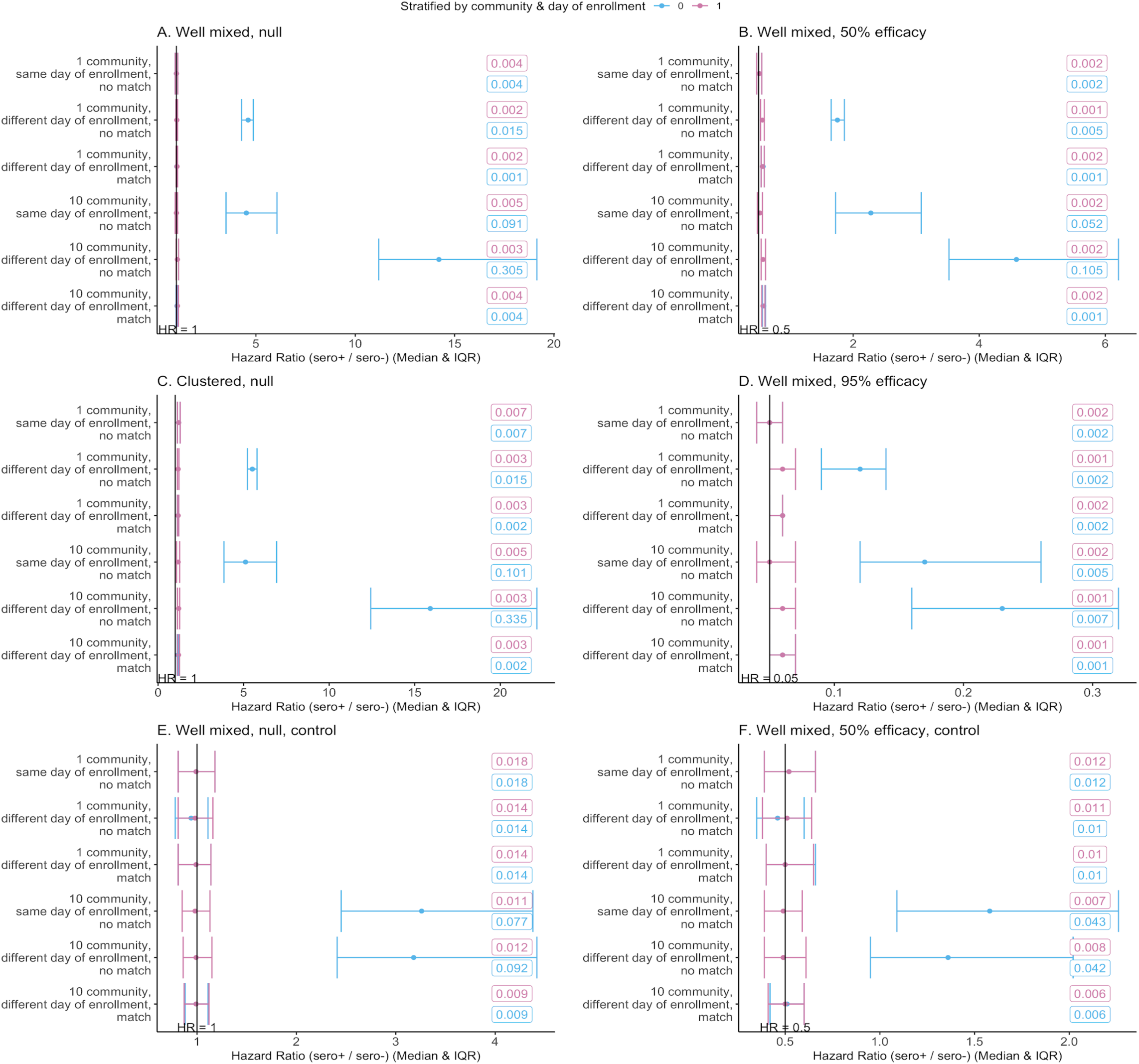
The median and IQR of simulations for each set of parameters are shown. Standard errors are reported in the boxes on each graph. Note the different x-axis scales. Simulations with zero events in either the seropositive or seronegative arm or with a hazard ratio >100 were excluded (percent of simulations excluded in each figure: A: 0.03%, B: 0.07%, C: 0.07%, D: 0.97%, E: 3.00%, F: 6.02%). For analyses with a high infection hazard for any enrolled individuals (e.g., Figures 1B and 1D with different days of enrollment), the estimated hazard ratio is between the ratio of the force of infection between seropositive and seronegatives (*β*^+^*/β^−^*) and the null HR=1. For a high infection hazard, the hazard ratio is closer to the null than the ratio of the force of infection since an individual’s hazard is not simply the product of their number of contacts and the force of infection. This is not a bias in the conventional sense, but rather a difference between the ratio *β*^+^*/β^−^* and the parameter that is estimated by the Cox model.

Simulations are in well mixed communities, meaning everyone within a community is connected to each other, except in Figure 1C which has clustering within communities. This clustering leads to correlations between infection status of particular individuals close together in the network and may be understood as creating multiple smaller (albeit overlapping) “communities” within each discrete community.

For simulations with one well mixed community with the same day of enrollment for all individuals (top lines of Figures 1A,1B, and 1D), a crude analysis returns unbiased results. If enrollment occurs on different days (Figures 1A,1B, and 1D, second and third lines), a crude analysis yields an upwardly biased estimate of the hazard ratio, making seropositivity appear harmful. However, matching or stratifying the analysis by day of enrollment removes this bias.

With multiple communities (and thus multiple, unconnected epidemics, Figures 1A,1B, and 1D, bottom halves), an unadjusted analysis creates the same upward bias, regardless of whether enrollment is on the same or multiple calendar dates, as the same calendar date does not mean the same phase of the epidemic in each of the communities. Once again, the bias is upward because individuals in communities with larger or more advanced epidemics are exposed to higher hazards and are more likely to be seropositive at baseline (Figures 2A-D). As before, the bias can be removed by stratifying and/or matching, this time on both community and day of enrollment. For analyses with a high infection hazard for any enrolled individuals (e.g., Figures 1B and 1D with different days of enrollment), the estimated hazard ratio is between the ratio *β*^+^/*β*^−^ and the null HR=1. For a high infection hazard, the hazard ratio is closer to the null than the ratio of the force of infection since an individual’s hazard is not simply the product of their number of contacts and the force of infection. This is not a bias in the conventional sense, but rather a difference between the ratio *β*^+^/*β*^−^ and the parameter that is estimated by the Cox model. For settings with a lower force of infection or fewer infectious contacts, this difference is imperceptible.

**Figure 2.**
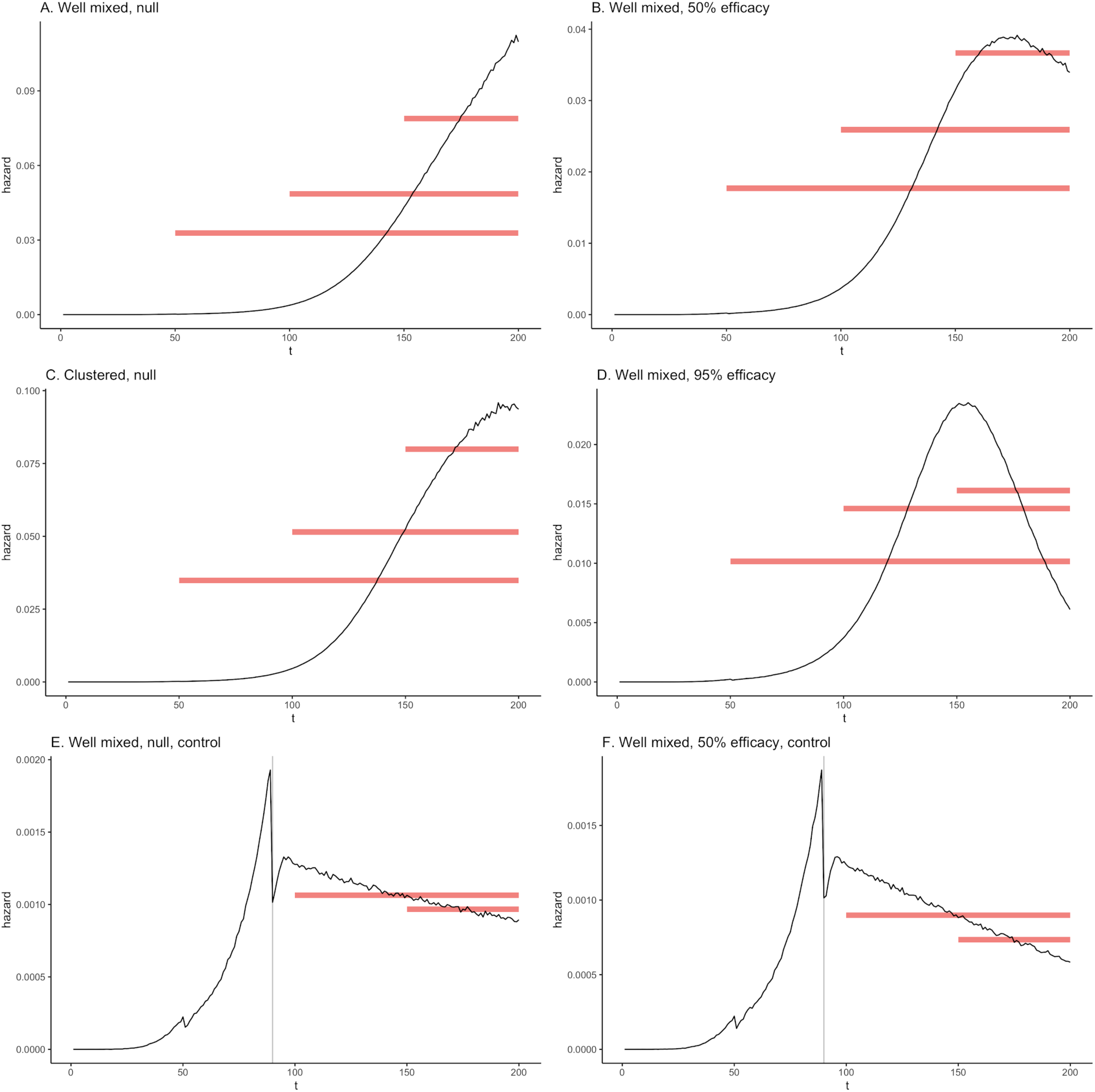
The average simulated daily hazard of infection for those in the initial susceptible compartment (i.e. never infected) to move to the exposed compartment in the simulations with one community. Note the different y-axis scales. Red bars show lengths of follow-up for each day of enrollment. The height of the bars indicates the average hazard for that duration of follow-up. In A-D, follow-up begins on days 50, 100, and 150 while in E and F, follow-up begins on days 100 and 150 only. Grey lines denote day control measures are implemented, which reduce the force of infection by 50% (E and F). The number of infectious individuals continues to grow beyond the day of control for approximately the average length of the latent period (5.6 days) due to those infected in the days just before control. This causes the hazard to increase again after its initial drop before declining again.

Clustering of contacts within communities (a departure from the assumption of a well mixed epidemic) produces an upward bias even in the matched / stratified analyses. As noted, this reflects that the different parts of the network have different local prevalence at any given time, resulting in a milder form of the same heterogeneity seen when comparing one to many communities. Because these clusters of high and low prevalence areas overlap and arise during the study, there is no a priori way to adjust for them.

In the simulations summarized in Figures 1E and 1F, transmission is reduced partway through the outbreak in one or more well mixed communities, reflecting control measures. As before, the single-community estimates are unbiased when all individuals enroll on the same day, but when enrollment occurs on different days or there are multiple communities, the estimates are biased. In the single-community simulations with two different days of enrollment, the unstratified, non-matched analysis estimates are slightly biased away from the null, making seropositivity look protective. This occurs because there are more seropositives at later enrollment dates when the average hazard over the rest of the study is lowest (Figures 2E and 2F). Hence, confounding can go in either direction depending on the dynamics of the epidemic at the times of enrollment. Matching alleviates the different biases, as does stratification in cases where there are infections in both the seropositive and seronegative arms. If there are substantially fewer seropositive individuals than seronegative individuals and the risk of infection after enrollment is low (i.e., because of effective control measures), there can be settings with no infections among the seropositive enrollees in some or all strata. In these cases, stratified analyses can lead to unstable results. Matched analyses are thus preferable because they remove this imbalance between the two exposure arms.

We note that in the simulations under the null with no control measures (Figures 2A and 2C), the daily hazard (proportion in the S compartment moving to the E compartment) initially increases during the early spread of the virus and then begins to plateau. In simulations with control and/or immunity (Figures 2B, 2D–F), the daily hazard increases and then decreases.

## DISCUSSION

We find that in studies assessing the efficacy of serostatus on future infection, comparing seropositive individuals to seronegative individuals with similar time-dependent patterns of exposure to infection is essential, because otherwise confounding can bias results: accounting for differential exposure among seropositive individuals and seronegative individuals is necessary to prevent bias. This bias can arise from either having multiple days of enrollment over the course of the study by design or by having multiple communities where the outbreak stochastically starts at different times. Stratifying or matching on community and day of enrollment alleviates this bias in well mixed communities. When there is clustering within communities, a slight upward bias remains, suggesting the local network structure in a trial is an important factor to consider.

While most individuals are no longer infectious when they seroconvert, it is possible for individuals to still be infectious (and thus not at risk for infection) when they seroconvert. Conversely, it is also possible for individuals to have already been reinfected when they seroconvert. For potentially asymptomatic infections, these cases would not be able to be excluded in a trial without viral testing. Excluding individuals who are infected soon after enrollment (e.g., within the average latent period length) would remove many of these cases. Additionally, because seroconversion is assumed to be detectable seven days after the end of the latent period and the average length of infectiousness is five days, individuals may move into the S’ compartment, where their susceptibility to infection is reduced in scenarios with seroprotection, before they are detectable as seropositive. For individuals enrolled into the study during this short window, they will be misclassified as seronegative (i.e., imperfect sensitivity), which could bias results towards the null.

The results shown here assume perfect specificity of the serologic test. Imperfect specificity will cause the usual biases due to misclassification (12), but may also exacerbate biases shown here, since there will be more false positives at low prevalence than at high prevalence, leading to differential misclassification depending on enrollment date. More complex interactions of immunity and infection, including immunity that wanes over the time scale of the study, viral-load dependent infection, and effects of repeated exposures, such as boosting of titers, may affect these biases as well, or introduce other potential biases. Further research is needed to understand the effects of these biological mechanisms in the specific context of SARS-CoV-2.

These simulations focus on the bias inherent in some study designs that may be considered, but do not address the feasibility of implementing these designs. In addition, we do not focus on the power of these studies; this may have important consequences in determining an adequate sample size. Sample size considerations will be particularly important in balancing the advantage of starting enrollment later, when the cumulative incidence is higher and thus the exposure arms are more likely to be balanced, and avoiding the tail of an outbreak or a setting after control measures have been implemented, which will reduce the infection risk for all participants. We have shown that matching can address these issues, but further work is needed to assess feasibility and sample size issues.

As serologic studies begin, understanding potential sources of bias and how to alleviate them are important for accurately estimating the extent and duration of immunity to SARS-CoV-2. Here we have focused on the impact of epidemic dynamics on estimation of seroprotection and have assumed all individuals in the model are exchangeable and differ only in whom they contact. Future work could examine additional heterogeneity, such as behaviors or factors that increase risk of infection, which might lead to further biases.

## Data Availability

Code will be available on github.

https://github.com/rek160/serologic-studies

## Appendix 1: Data generating details - network and outbreak

### Generate a network

We use a stochastic block model to generate a network graph, using the sample_sbm function in the R package igraph (14). In our simulations, we create networks with one single community and networks with 10 communities. We keep the total population across simulations constant at 10,000. Therefore in simulations with 1 community, there are 10,000 nodes in that one community, and for simulations with 10 communities, there are 1,000 people in each community.

The sample_sbm function conducts a Bernoulli trial for each potential edge in the graph. In our “well mixed” communities, the probability for each edge within the same community is 1, meaning all nodes in the community are connected. The probability of connection for nodes in different communities is 0, meaning there are no edges between communities. For “clustered” communities, the probability of an edge between nodes in different communities remains 0. While in the well mixed communities, the probability of an edge between nodes in the same community was 1, here it is greatly reduced. For the single community simulations, the probability of an edge is 0.002, and for the 10 community simulations, the probability of a within community edge is 0.02, meaning the expected number of edges for each node is approximately 20. This creates smaller, overlapping communities, or clusters, within each larger discrete community.

In order to keep R_0_ constant (1.5) in all simulations, we use the following formula (4,13) to calculate *β* the force of infection):

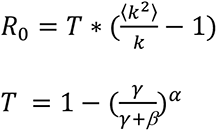

where k is the mean degree of the network, γ is the infectious period rate, *α* is the infectious period shape.

### Seeding outbreak

The outbreak in the communities is seeded by introductions from an outbreak in an external population (9) of one million individuals. All introductions occur between days 1 and 50. The number of nodes infected externally on a given day is based on a binomial distribution, with the probability equal to 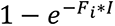 where F_i_ is the proportionality constant for the amount of contact between the external population and a node in community *i*, and I is the number of infected individuals in the external population, which has an exponentially growing deterministic outbreak from day 1 to day 50. 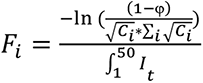 where φ = expected number of introductions, C_i_ = size of community i and It is the number of infected individuals in the external population on day t. The probability of introduction for a given node scales with the size of that node’s community; however in our simulations, all communities have the same size so the probability is equal.

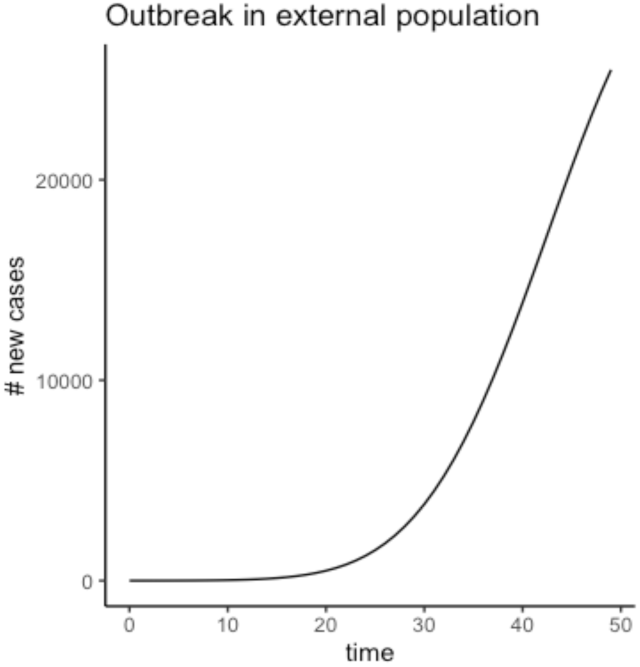

### Outbreak in communities

When a node is infected, either from the external population or from an infected node in the community, they move from the susceptible compartment to the exposed compartment. Their latent period, the time between exposure and onset of infectiousness, is drawn from a gamma distribution with mean 5.6 days, independent of the period for any other node. After the latent period ends, individuals progress to the infectious compartment. Their infectious period is drawn from a gamma distribution with a mean of 5 days, independent of the period for any other node. After their infectious period ends, they move into the susceptible’ compartment.

On each day, an infectious node has a daily probability of infecting each of the susceptible (seronegative) nodes they are connected to of 1 − *e*^−^*β*^−^ where *β*^−^ is the force of infection for those initially susceptible, and a daily probability of infecting each of the susceptible’ (seropositive) nodes they are connected to of 1 − *e*^−^*β^+^* where *β*^+^ is the force of infection for those who have been infected previously. The nodes they infect then move into the exposed compartment and the steps above repeat.

